# Workplace Pollution and Risk of Incident Coronary Artery Disease

**DOI:** 10.64898/2026.01.16.26344285

**Authors:** Hua Gao, Kai-Uwe Jarr, Yoko Kojima, Ting Xiong, Juyong Brian Kim, Paul Cheng, Leonardo Trasande, Annette Peters, Gerard Pasterkamp, Chiara Giannarelli, Nicholas J. Leeper

## Abstract

**Importance:** Genetic factors only explain ∼50% of an individual’s lifetime CAD risk. Workplace pollution exposure likely represents a significant, under-recognized, and modifiable contributor. Clarifying its independent effect – distinct from residential air pollution – could inform targeted prevention, clinical risk assessment, and policy strategies.

**Objective:** To quantify the independent association between workplace pollution exposure and incident CAD, rigorously adjusting for canonical risk factors, genetic risk, socioeconomic deprivation index, and residential air pollution.

**Design, Setting, and Participants:** A prospective cohort analysis of 103,599 adults in the UK Biobank with complete baseline employment history and specialized workplace environment surveys. Participants with prior CAD, or missing genetic information, canonical risk factor data, or residential air pollution measurements, were excluded.

**Exposure:** Cumulative, self-reported duration of exposure to workplace pollutants (including dust, smoke, exhaust, chemicals, asbestos, paints, and pesticides), summarized as a percentile measure.

**Main Outcomes and Measures:** Incident CAD. Associations were estimated using Cox proportional hazards models adjusted for demographics, comorbidities, CAD polygenic risk score, and residential air pollution metrics, accounting for competing death events.

**Results:** The cohort (median age 64 years; 43% male) had high baseline prevalence of smoking (41%) and co-morbidities (hypertension 27%, hyperlipidemia 16%, diabetes 4.3%). Common exposures included smoke (56%), dust (39%), and chemicals (27%). Over a median 7.5-year follow-up, 4,327 CAD cases occurred. Compared to the low exposure group, high workplace pollution exposures showed a stronger unadjusted association with CAD (hazard ratio [HR], 1.51; 95% CI, 1.40-1.63) relative to residential air pollution (unadjusted HR, 1.23; 95% CI, 1.14-1.33). Importantly, in multivariable models, workplace pollution remained independently associated with higher CAD incidence (adjusted HR, 1.21; 95% CI, 1.03-1.41), robust to all adjustment. Significant specific drivers included paints, thinners or glues, diesel exhaust, fumes, asbestos, and dust.

**Conclusions and Relevance:** Workplace pollution is confirmed as a robust, independent risk factor for incident CAD. Its association is demonstrably stronger than residential air pollution and persists despite comprehensive adjustment for genetic predisposition and traditional cardiovascular risk factors. These findings underscore the need to strengthen worker protections, integrate occupational history into clinical risk assessment, and prioritize mechanistic research into non-lipid pollution pathways.

## Introduction

Cardiovascular disease remains the leading global cause of mortality^1^. While genetic predisposition accounts for approximately 50% of coronary artery disease (CAD) susceptibility^2,3^, a substantial proportion of lifetime risk remains unexplained. Environmental exposures, particularly to inhalable pollution, are hypothesized to drive part of this residual risk. Although residential air pollution is a well-established risk factor^4–9^, occupational pollution exposure represents an understudied, potentially modifiable source of concentrated exposure that is not routinely integrated into clinical risk assessment. Workers in occupational settings involving smoke, exhaust, dust, chemicals, and industrial materials frequently experience high-intensity, chronic exposures substantially greater than typical residential levels. Therefore, quantifying the independent contribution of workplace pollution to incident CAD risk – while robustly accounting for both canonical risk factors and genetics – is critical. Existing evidence is limited: several retrospective studies which did not fully adjust for conventional risk factors implied certain occupational exposures are associated with increased CAD mortality^10,11^; however, comprehensive prospective data are lacking. To address this critical knowledge gap, we conducted a large cohort study using the UK Biobank to characterize the independent impact of cumulative workplace pollution exposure on incident CAD.

## Methods

### Study Design and Population

UK Biobank has collected extensive health, genetic, and lifestyle data from over 500,000 anonymous volunteers in the UK. We focused on the 120,075 participants who completed work environment questionnaires. The enrollment date was defined as the survey completion date.

Exclusion criteria included: prevalent CAD diagnosis prior to the enrollment date; missing genetic information or conventional risk factor data or deprivation index; and missing residential air pollution measurements. The follow-up extended to latest update or death until this study conduct in Sep 2025.

The UK Biobank fields and coding strategies used to construct the cohort are detailed in eMethods.

### Exposure

In the survey, participants completed an occupational history spanning multiple employment periods. For each period, they reported whether their workplace had dust; chemical or other fumes; cigarette smoke; materials containing asbestos; paints, thinners or glues; pesticides; or diesel exhaust. From participants’ responses, we calculated cumulative exposure duration (years) and transformed it into percentile scores (ranging from 0 to 1). The overall composite workplace pollution metric was defined as the maximum percentile score across all seven components.

### Outcome and covariates

Incident CAD was ascertained via linked health records. The incident event date was taken as the earliest occurrence across any linked source.

We adjusted for canonical CAD risk factors (age, sex, ethnicity, smoking history, prevalent diabetes, hyperlipidemia, and hypertension), genetic predisposition proxied by CAD polygenic risk score, socioeconomic deprivation index, and residential air pollution. Each residential air pollution component (NO2, NOx, PM10, PM2.5, and PM2.5-10) was similarly transformed into percentiles, and the overall residential air pollution was also summarized as the maximum across all components.

### Statistical analysis

To examine etiologic associations between exposures and incident CAD, we fitted a cause-specific Cox’s proportional hazard model, treating death as a competing event and adjusting for all the covariates and exposures listed above. Hazard ratios (HRs) are reported per one unit increase in the percentile scale, representing a change from the lowest (0%) to highest (100%) level of exposure. Sensitivity analyses were performed by substituting the composite residential air pollution with the original or percentile-transformed PM2.5 and/or PM10, and by replacing the composite workplace pollution with individual components.

## Results

### Baseline characteristics

A total of 103,599 participants were analyzed (Figure 1A, Table 1; median age 64 years; 43% male; 41% ever smokers; and 98% White). Prevalent cardiometabolic conditions included hypertension (27%), hyperlipidemia (16%), and diabetes (4.3%).

**Figure 1.**
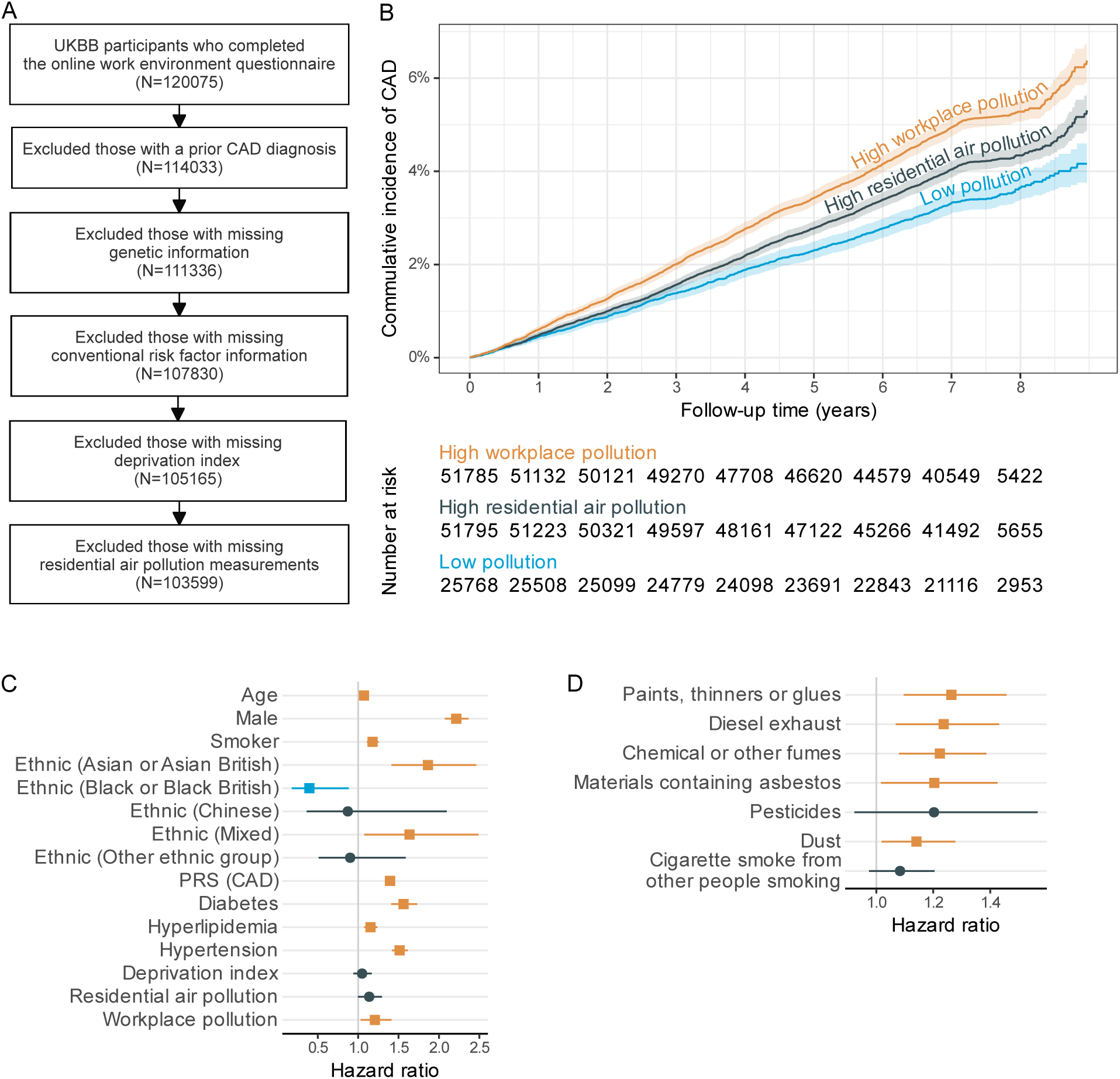
Cohort design and associations of workplace pollution with incident CAD. A. Flow diagram of UK Biobank participant inclusion and exclusion for the prospective cohort. B. Cumulative incidence curves for incident CAD comparing high pollution groups versus low pollution group. High/low defined by median splits of the respective composite percentile-based metrics. Death treated as a competing event. C. Forest plot of adjusted cause-specific Cox proportional hazards models for incident CAD. PRS stands for polygenic risk score. D. Forest plot of adjusted associations for individual workplace exposure components. Error bars indicate 95% confidence intervals.

**Table 1.** Baseline characteristics of the analytic cohort.

| **Characteristic** | **N = 103,599** |
| --- | --- |
| Age | 64 (57, 69) |
| Male | 44,354 (43%) |
| Smoker | 42,484 (41%) |
| Ethnic |  |
| White | 101,329 (98%) |
| Asian or Asian British | 660 (0.6%) |
| Black or Black British | 516 (0.5%) |
| Chinese | 219 (0.2%) |
| Mixed | 463 (0.4%) |
| Other ethnic groups | 415 (0.4%) |
| Polygenic risk score (CAD) | -0.21 (-0.86, 0.42) |
| Diabetes | 4,446 (4.3%) |
| Hyperlipidemia | 16,104 (16%) |
| Hypertension | 27,696 (27%) |
| Deprivation index |  |
| England | 11.0 (6.5, 19.1) |
| Scotland | 6.2 (3.5, 13.1) |
| Wales | 8.7 (5.5, 15.2) |
| Workplace pollution (years of exposure) |  |
| Cigarette smoke from other people smoking | 56% (17 [8, 34]) |
| Dust | 39% (19 [7, 34]) |
| Chemical or other fumes | 27% (15 [6, 32]) |
| Paints, thinners or glues | 17% (16 [6, 33]) |
| Diesel exhaust | 16% (17 [6, 34]) |
| Materials containing asbestos | 9.5% (19 [8, 36]) |
| Pesticides | 4.0% (12 [5, 27]) |
| Residential air pollution for the year of 2010 ($\mu\text{g}/\text{m}^3$) | |
| NO<sub>2</sub> | 25 (21, 31) |
| NO<sub>x</sub> | 41 (33, 50) |
| PM<sub>2.5</sub> | 9.84 (9.18, 10.46) |
| PM<sub>10</sub> | 15.97 (15.14, 16.93) |
| PM<sub>2.5-10</sub> | 6.09 (5.83, 6.59) |

Continuous variables are shown as median (interquartile range [IQR]);
Categorical variables as n (%);
Workplace pollutions are presented as % ever exposed (exposed median [exposed IQR]).

Workplace pollution exposures were common (Table 1, eFigure 1): 56% reported smoke exposure at the workplace (median duration among exposed: 17 years), 39% dust (19 years), 27% chemical fumes (15 years), 17% paints/thinners/glues (16 years), 16% diesel exhaust (17 years), 9.5% asbestos (19 years), and 4.0% pesticides (12 years).

### Higher CAD incidence in high workplace pollution group

Over a median follow-up of 7.5 years, 4,327 incident CAD events were recorded (Figure 1B). In the reference group (low workplace and residential air pollution group, N = 25,768), 870 events occurred. High residential air pollution (N = 51,795; 2,136 events) was associated with an unadjusted HR of 1.23 (95% CI, 1.14-1.33). However, the high workplace pollution group (N = 51,785; 2,598 events) demonstrated a notably larger unadjusted HR of 1.51 (95% CI, 1.40-1.63), indicating a stronger crude association for workplace pollution exposure than residential air pollution exposure.

### Workplace pollution as an independent CAD risk

In the fully adjusted model, residential air pollution demonstrated a borderline significant association with CAD, with an adjusted HR of 1.14 (95% CI, 1.00-1.29) (Figure 1C).

Crucially, workplace pollution remained significantly independently associated with higher CAD incidence. The adjusted HR was 1.21 (95% CI, 1.03-1.41).

When analyzing individual workplace components, all components yielded positive hazard ratios (Figure 1D, eTable 1). Statistically significant associations were observed for exposure to paints/thinners/glues (HR 1.26; 95% CI, 1.10-1.46), diesel exhaust (1.24; 1.07-1.43), chemical or other fumes (1.22; 1.08-1.39), asbestos (1.20; 1.02-1.43), and dust (1.14; 1.02-1.28).

## Discussion

This large prospective cohort study showed that exposure to workplace pollution was associated with a higher incidence of CAD, independent of conventional risk factors and genetic predisposition. Importantly, the association was also independent of and notably larger than the risk associated with residential air pollution. Multivariable modeling suggests that minimizing workplace pollution exposure could reduce the relative 5-year risk for CAD by 4.3% (eFigure 2), highlighting the need to integrate occupational exposure assessments into cardiovascular prevention efforts and the potential benefit of enhancing protections for high-risk jobs.

Most mechanistic CVD research has centered on traditional risk factors and heritable pathways related to lipid handling, inflammation, vascular remodeling, and thrombosis. However, these collectively account for only a portion of lifetime risk, underscoring environmental exposures as underappreciated drivers of vascular disease. Robust epidemiologic and mechanistic data have linked ambient air pollution to higher rates of cardiovascular morbidity and mortality plausibly through systemic inflammation and oxidative stress, endothelial dysfunction, autonomic imbalance, and prothrombotic signaling^7^. Occupational exposures likely add an additional, distinct layer beyond traditional risk factors, genetics, and even residential ambient air. Notably, the elevated risk persisted regardless of underlying hyperlipidemia status (eFigure 3), supporting the concept that environmental toxins may mediate disease via non-lipid-centric pathways. Together, these observations reinforce the need to systematically interrogate the occupational and broader “exposome-related” risk factors and their effects on vascular biology.

This study has limitations. First, exposure to workplace pollution was self-reported and may be subject to recall bias and reporting variability. Although we harmonized responses, objective measurements would improve precision. Second, due to the rarity of biobanks with detailed workplace pollution data, we could not replicate these findings in an independent cohort. Therefore, generalizability may be limited.

## Conclusions

Our findings confirm workplace pollution exposure is an independent risk factor for incident CAD, persisting after adjustment for genetic and traditional risk factors. Notably, this association proved stronger than residential air pollution. Significant drivers included paints/thinners/glues, diesel exhaust, chemical or other fumes, asbestos, and dust. These results underscore the need to integrate occupational exposure history into cardiovascular prevention strategies, coupled with strengthened workplace protections for high-exposure workers. Finally, we highlight the urgency of advancing mechanistic research to delineate the non-lipid pathways though which these environmental pollutants drive CAD.

## Supporting information

Supplemental Online Content

## Data Availability

All data are available online at UKBB upon approval.

## Acknowledgments

This research has been conducted using the UK Biobank Resource under application number 706110.

## Author Contributions

Hua Gao and Nicholas Leeper had full access to all of the data in the study and takes responsibility for the integrity of the data and the accuracy of the data analysis.

*Concept and design*: Nicholas Leeper, Hua Gao, Kai-Uwe Jarr

*Acquisition, analysis, or interpretation of data*: Hua Gao, Nicholas Leeper

*Drafting of the manuscript*: Hua Gao, Kai-Uwe Jarr, Nicholas Leeper

*Critical review of the manuscript for important intellectual content*: all authors

*Statistical analysis*: Hua Gao

*Obtained funding*: Nicholas Leeper

*Supervision*: Nicholas Leeper

