## Supplemental Online Content for "Workplace Pollution and Risk of Incident Coronary Artery Disease"

**eFigure 1**. Distribution and correlation of continuous covariates at baseline. A. Distribution of the original and percentile-transformed combined deprivation index. B. Distribution of the original and percentile-transformed residential air pollution metrics. C. Distribution of the original and percentile-transformed workplace pollution metrics. D. Distribution of the composite residential air pollution and workplace pollution metrics. E. Correlation matrix of the deprivation index, residential air pollution and workplace pollution.
**eFigure 2**. Observed cumulative incidence and estimated counter-factual cumulative incidence curve under highest or lowest workplace pollution exposure.
**eFigure 3**. Cumulative incidence curves for incident CAD among participants with high workplace pollution exposure, stratified by hyperlipidemia comorbidity status.

**eTable 1**. Sensitivity analyses substituting alternative residential air pollution and workplace pollution metrics.

**eMethods**

1. UK Biobank fields in use and coding strategies.
2. Survival model adjustment and sensitivity analyses.

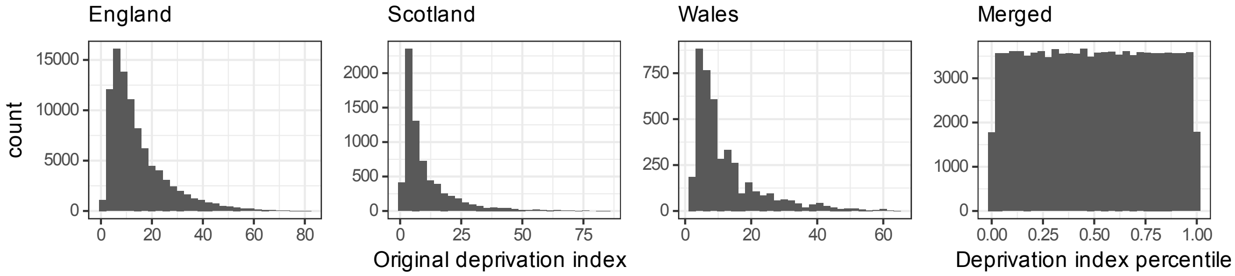


**eFigure 1**. Distribution of continuous covariates at baseline. A. Distribution of the original and percentile-transformed combined deprivation index.

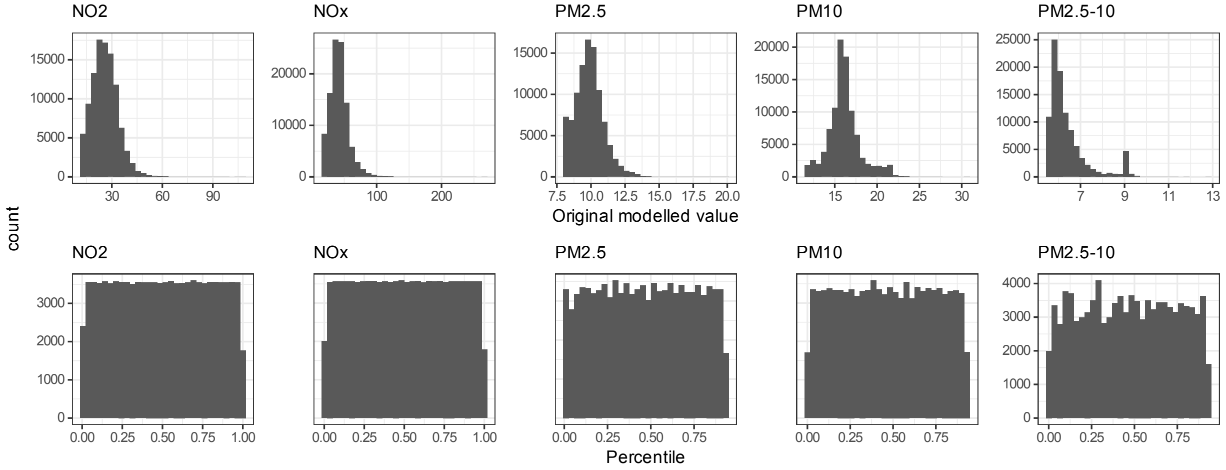


**eFigure 1**. Distribution of continuous covariates at baseline. B. Distribution of the original and percentile-transformed residential air pollution metrics.

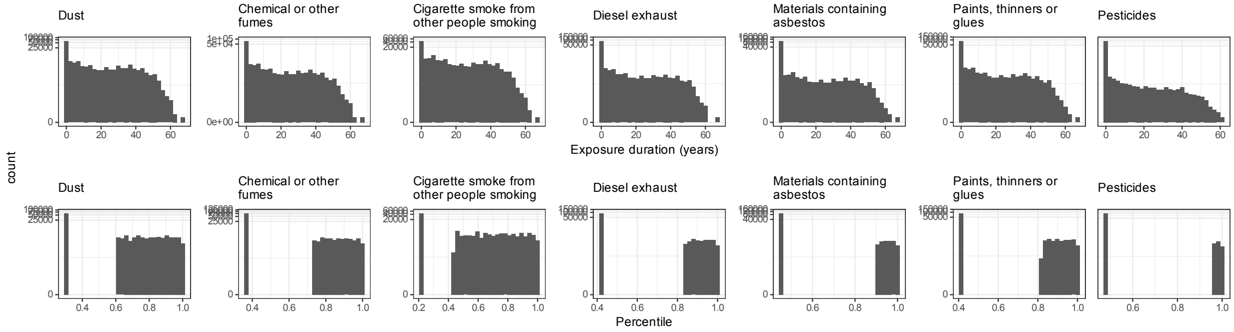


**eFigure 1**. Distribution of continuous covariates at baseline. C. Distribution of the original and percentile-transformed workplace pollution metrics.

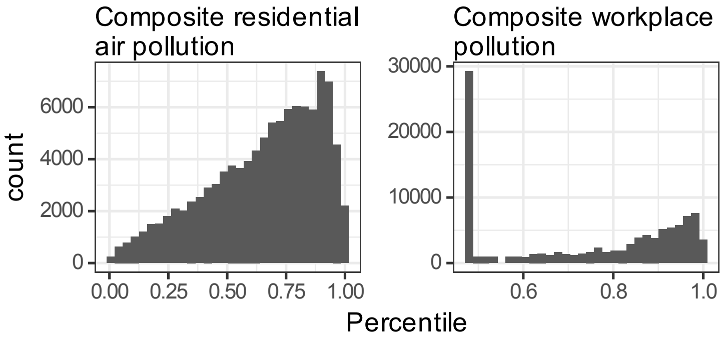


**eFigure 1**. Distribution of continuous covariates at baseline. D. Distribution of the composite residential air pollution and workplace pollution metrics.

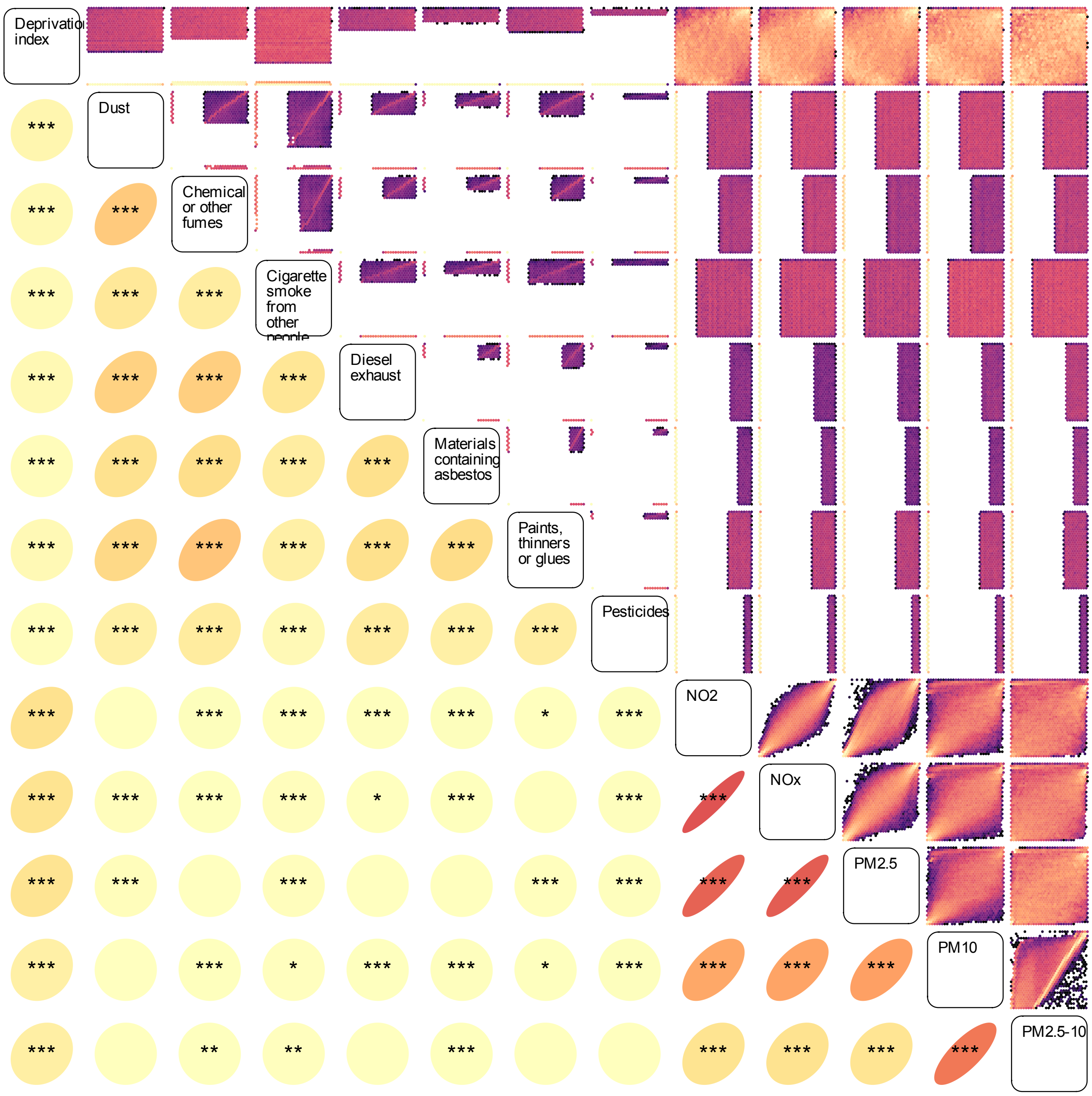


**eFigure 1**. Distribution of continuous covariates at baseline. E. Correlation matrix of the deprivation index, residential air pollution and workplace pollution. Ellipses at the lower triangle present the Spearman correlation coefficients. Asterisks denote statistical significance: "*" for *P*-value <= 0.05, "**" for 0.01, and "***" for 0.001.

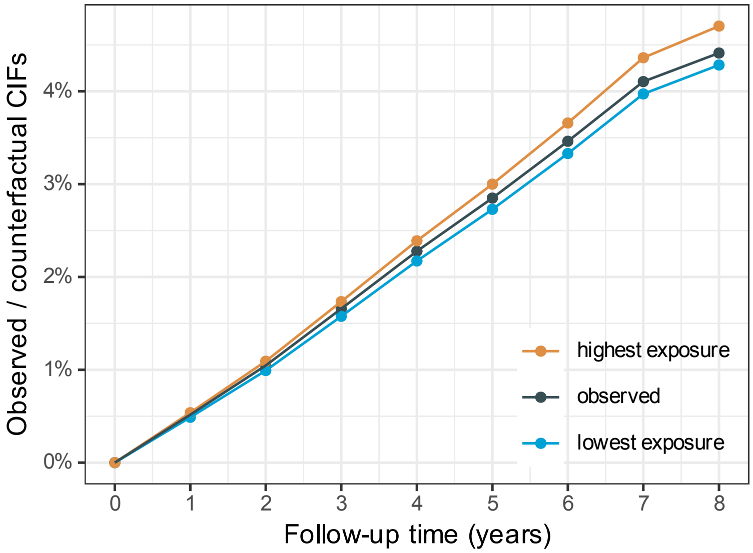


**eFigure 2**. Observed cumulative incidence and estimated counter-factual cumulative incidence curve under highest or lowest workplace pollution.

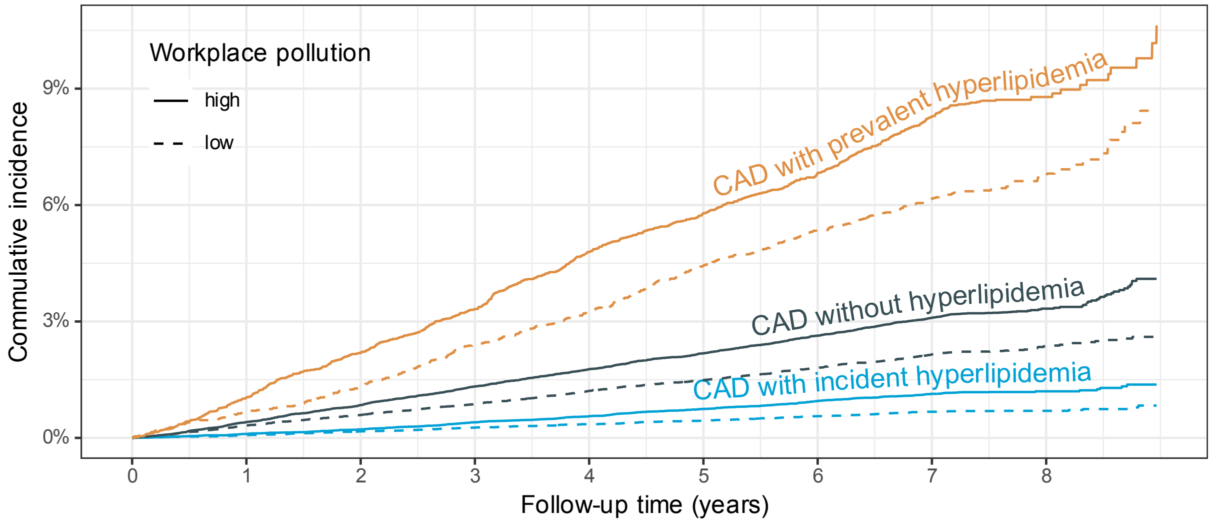


**# at risk**

| | | | | | | | | |
|---|---|---|---|---|---|---|---|---|
| CAD with prevalent hyperlipidemia | | | | | | | | |
| 9096 | 8951 | 8745 | 8549 | 8191 | 7944 | 7567 | 6792 | 964 |
| 7008 | 6925 | 6792 | 6651 | 6442 | 6254 | 5965 | 5457 | 871 |
| CAD without hyperlipidemia | | | | | | | | |
| 42689 | 42181 | 41376 | 40721 | 39517 | 38676 | 37012 | 33757 | 4458 |
| 44806 | 44377 | 43690 | 43199 | 42089 | 41382 | 39959 | 36972 | 5029 |
| CAD with incident hyperlipidemia | | | | | | | | |
| 42689 | 42181 | 41376 | 40721 | 39517 | 38676 | 37012 | 33757 | 4458 |
| 44806 | 44377 | 43690 | 43199 | 42089 | 41382 | 39959 | 36972 | 5029 |

**Cum. events**

| | | | | | | | | |
|---|---|---|---|---|---|---|---|---|
| CAD with prevalent hyperlipidemia | | | | | | | | |
| 0 | 95 | 198 | 300 | 431 | 517 | 607 | 725 | 757 |
| 0 | 46 | 90 | 168 | 224 | 304 | 364 | 416 | 435 |
| CAD without hyperlipidemia | | | | | | | | |
| 0 | 174 | 360 | 560 | 746 | 912 | 1095 | 1269 | 1324 |
| 0 | 143 | 263 | 387 | 536 | 657 | 791 | 930 | 972 |
| CAD with incident hyperlipidemia | | | | | | | | |
| 0 | 44 | 93 | 170 | 236 | 311 | 394 | 464 | 481 |
| 0 | 32 | 75 | 117 | 156 | 196 | 245 | 291 | 299 |

**eFigure 3**. Cumulative incidence curves for incident CAD among participants with high workplace pollution exposure, stratified by hyperlipidemia comorbidity status. “Prevalent” indicates diagnosis before enrollment; “Incident” indicates diagnosis after enrollment.

**eMethods**

**1. UK Biobank fields in use and coding strategies**

A. Exposure assessment

In the survey, participants completed an occupational history spanning multiple employment periods (UKBB category 123). For each period, they reported whether their workplace: was very dusty (Field 22609), was full of chemical or other fumes (Field 22610), had a lot of cigarette smoke from other people smoking (Field 22611), involved working with materials containing asbestos (Field 22612), involved working with paints, thinners or glues (Field 22613), involved working with pesticides (Field 22614), or had a lot of diesel exhaust (Field 22615). Response options included "often", "sometimes", "rarely/never", and "do not know". For binary classification, "often" and "sometimes" were classified as exposed, "rarely/never" as unexposed, and "do not know" as missing. For each exposure component, we summed the duration (in years) across all employment periods classified as exposed to derive cumulative years of exposure.

To mitigate the influence of skewed distribution and extreme values, each component's cumulative duration was transformed into within-cohort percentile scores (ranging from 0 to 1). The overall composite workplace pollution metric was defined as the maximum percentile score across all seven specific exposure components, thereby capturing the dimension of dominant or peak workplace exposure. For estimating unadjusted cumulative incidence curves, this composite metric was dichotomized at the cohort median into low (median and below median) versus high exposure (above median) exposure groups.

B. Outcomes

Incident CAD was ascertained via linked health records within the biobank, by combining all hospital inpatient diagnoses (Category 2002) with the harmonized "first occurrences" dataset (Category 1712), which aggregates primary care data, hospital inpatient data, death register records, and self-reported medical condition codes. CAD events were defined using ICD-10 codes: I21 (acute myocardial infarction), I22 (subsequent myocardial infarction), I23 (certain current complications following acute myocardial infarction), I24 (other acute ischemic heart diseases), and I25 (chronic ischemic heart disease), but excluding I25.3 (aneurysm of heart) and I25.4 (coronary artery aneurysm). The incident event date was taken as the earliest occurrence across any linked source. Participants with evidence of CAD prior to enrollment were excluded from the analysis.

C. Covariates

We adjusted for canonical CAD risk factors including age, sex (Field 31), ethnicity (Field 21000), smoking history, prevalent diabetes, hyperlipidemia, and hypertension. Age at enrollment was calculated from the enrollment date, i.e. the survey completion date (Field 22500), and birth year/month (Field 34 and 52). Smoking history was obtained from a series of online follow-up surveys (Field 20116) and was dichotomized as ever-smoker versus never-smoker at enrollment. The ICD-10 definitions for prevalent cardiometabolic conditions were diabetes (E08-E11 and E13-E14), hyperlipidemia (E78 and E88.8 but excluding E78.7 and E78.8) and hypertension (I10, I12, I15).

Besides the canonical CAD risk factors, we also adjusted for genetic predisposition to CAD, socioeconomic deprivation index and residential air pollution.

Genetic predisposition to CAD was proxied by the CAD polygenic risk score (Field 26227), derived by meta-analyzing three non-overlapping external GWAS sources.

The socioeconomic deprivation index (Category 76) comes from a UK government qualitative study of deprived areas in British local councils. The study is conducted separately in England, Scotland and Wales. As the scores are calculated in different ways between England (Field 26410), Scotland (Field 26427) and Wales (Field 26426), it's not advisable to directly combine the original values. Thus, we transformed the deprivation index into percentile scaled from 0 to 1 in each region before combining.

As a comparator for workplace pollution exposure, residential air pollution was characterized using estimates (Category 114) for the year 2010, which were modeled for each postcode address using a Land Use Regression model based on monitoring data from European Study of Cohorts for Air Pollution Effects. Components included nitrogen dioxide (Field 24003), nitrogen oxides (Field 24004), PM10 (Field 24005), PM2.5 (Field 24006), and PM2.5-10 (Field 24008). Similarly, each component was transformed to percentile scaled from 0 to 1, and the overall residential air pollution was also summarized as the maximum of the component-specific percentiles. For cumulative incidence curve estimation, this composite residential air pollution metric was dichotomized at the cohort median.

The distribution and correlation of the exposure and covariates are shown in eFigure 1.

**2. Survival model adjustment and sensitivity analyses**

The cumulative incidence of CAD was estimated using a competing-risk framework with death treated as a competing event. We compared exposure groups (high workplace pollution or high residential air pollution) against the lowest-exposure reference group (low workplace and low residential air pollution).

To examine etiologic associations, we fitted cause-specific Cox's proportional hazard models for time to incident CAD, treating death as a competing event and adjusting for the covariates listed above. We evaluated both overall composite pollution metric and individual pollution components. Exposure variables were transformed to within-cohort percentiles across pollutants. Accordingly, hazard ratios (HRs) and 95% confidence intervals (CIs) are reported per one unit increase in the percentile scale, representing a change from the lowest (0%) to highest (100%) level of exposure.

Sensitivity analyses were performed by substituting the composite residential air pollution with the original or percentile-transformed PM2.5 and/or PM10, and by replacing the composite workplace pollution with individual components.
